# Meta-analysis of genotype-phenotype associations in Alström syndrome

**DOI:** 10.1101/2022.10.21.22281173

**Authors:** Brais Bea-Mascato, Diana Valverde

**Affiliations:** CINBIO, Universidad de Vigo, 36310, Spain; Grupo de Investigación en Enfermedades Raras y Medicina Pediátrica, Instituto de Investigación Sanitaria Galicia Sur (IIS Galicia Sur), SERGAS-UVIGO, Vigo, Spain

**Author notes:** Correspondence: Diana Valverde, CINBIO Facultad de Biología, Universidad de Vigo, Campus As Lagoas-Marcosende s/n, 36310 Vigo, Spain.

**Keywords:** ALMS, Alström syndrome, ciliopathy, genotype-phenotype, metabolic disease, obesity, meta-analysis, rare disease

## Abstract

**Introduction:** Alström syndrome (ALMS, #203800) is an ultra-rare monogenic recessive disease. This the syndrome is associated with mutations in the *ALMS1* gene, which codes for a centrosome structural protein responsible for centrosome cohesion. The type of mutation associated with ALMS is mostly cLOF (97%) and they are mainly located in exons 8, 10 and 16 of the gene. Other studies in the literature have tried to establish a genotype-phenotype correlation in this syndrome with little success. The difficulty in recruiting a large cohort in rare diseases is the main barrier to conducting this type of study.

**Methods:** In this study we have collected all cases published to date for ALMS. We have created a database with those patients who had a genetic diagnosis and an individualised clinical history. Finally, we have attempted to establish a genotype-phenotype correlation using the truncation site of the patient’s longest allele as a grouping criterion.

**Results:** We collected a total of 357 patients of which 227 had complete clinical information, complete genetic diagnosis and meta information regarding sex and age. We have seen that there are 5 variants in high frequency with p.(Arg2722Ter) being the most common variant with 28 alleles. There are no gender differences in disease progression. Finally, truncating mutations in exon 10 seem to be correlated with a higher prevalence of liver disorders in patients with ALMS.

**Conclusion:** The location of the mutation in the *ALMS1* gene does not have a major impact on the phenotype developed by the patient. Only mutations in exon 10 of the *ALMS1* gene were associated with a higher prevalence of liver disease.

## Introduction

Alström syndrome (ALMS, #203800) is an ultrarare monogenic disease caused by mutations in the *ALMS1* gene. It is an autosomal recessive disorder with an estimated incidence of 1-9 cases per-1,000,000 inhabitants and currently there are an estimated 1000 cases described worldwide (Orphanet; 03-05-2022).

Most of mutations associated with this disease are complete loss-of-function (cLOF) mutations that generate a stop codon either at the same site as the mutation or result in a frameshift alteration leading to a stop codon (Marshall et al., 2015). There are currently 388 loss-of-function variants reported in ClinVar and 253 in gnomAD. These pathogenic variants have a uniform distribution along the gene and are mainly located in coding regions. Whole exome sequencing (WEG) is a standard practice for the genetic testing of rare diseases, which implies that intronic regions are poorly studied, explaining why most pathogenic variants are detected in coding regions (Sawyer et al., 2016). Exons 8, 10 and 16 are considered mutation hotspots, but this seems to be due to their large size rather than a specific regulatory correlation. Exon 8 (6,108bp) for example covers 50% of the total gene sequence (12,844bp).

ALMS presents a very heterogeneous phenotype in which symptoms can be aggregates into two main groups (Marshall et al., 2007a, 2011). A first group includes the presence of retinal dystrophy from early life, various metabolic disorders (obesity, hypertriglyceridemia or type 2 diabetes mellitus), hearing loss, liver and kidney dysfunction and cardiac disorders such as dilated cardiomyopathy (DCM) (Marshall et al., 2005, 2007b, 2007a). The second group of symptoms would include short stature, recurrent pulmonary infections, mental and cognitive impairment and various endocrine disorders affecting the thyroid and reproductive system (Marshall et al., 2007a). This second group would also include other very rare symptoms, such as alterations in the fingers, alopecia and spinal abnormalities (Marshall et al., 2007a).

ALMS is characterised by high intra- and inter-familial phenotypic heterogeneity (Marshall et al., 2005, 2007a, 2007b, 2011). This means that siblings with the same genotype may have different phenotypes, which complicates the establishment of a genotype-phenotype correlation (Hollander et al., 2017).

Several studies have attempted to establish a genotype-phenotype correlation without success using cohorts with 12-18 patients (Bond et al., 2005; Minton et al., 2006). Studies in larger cohorts, 58 patients, have shown that there is correlation between disease-causing variants in exon 16 and the presence of retinal degeneration before one year of age, the occurrence of urological dysfunction, DCM and diabetes (Marshall et al., 2007b). Moreover, this study found a significant correlation between disease-causing variants in exon 8 and the absent, mild or delayed kidney disease.

Premature termination codon (PTC) mutations are often classified as cLOF and are associated with the activation of nonsense-mediated mRNA decay (NMD), which results in the elimination of the expression of the mutated gene (Supek et al., 2021). However, it has been shown that NMD is not 100% efficient when nonsense mutations are located in the last exons of genes (Kerr et al., 2001; Lindeboom et al., 2016; Supek et al., 2021). In the case of ALMS, there are patients who continue to express the ALMS1 protein even when carrying two cLOF variants (Chen et al., 2017). In that study of 23 patients where the clinical manifestations were related to the expression or non-expression of the *ALMS1* gene, it was observed that those patients with residual *ALMS1* gene expression presented milder phenotypes (Chen et al., 2017).

Although there are several databases that list the mutations described in the *ALMS1* gene (GnomAD, ClinVar), only one, LOVD, offers a register with 140 patients but the clinical data is absent or incomplete in many cases (Fokkema et al., 2011; Landrum et al., 2018; Karczewski et al., 2020). There is still no open patient registry that collects clinical and genetic information in a comprehensive way. In this study, we have reviewed 112 publications related to ALMS and compiled all available clinical and genetic information. We have collected a total of 357 patients, manually curated, on which various analyses have been performed to look for a genotype-phenotype correlation. Finally, we have made this dataset available to the scientific community in a single public access database (http://genesandfunctions.uvigo.es/alms-db/).

## Methods

### Preferred reporting items for systematic reviews and meta-analyses (PRISMA) guidelines

This meta-analysis was performed following the PRISMA guidelines, as described in Niederlova et al. (Niederlova et al., 2019), for Bardet Biedl syndrome (BBS). In this case, the main PICO question our study sought to address was: Do ALMS patients have different phenotypes depending on where in the gene, the stop mutation occurs?. Similar studies have been conducted in smaller cohorts grouping patients by major mutational hotspots in the *ALMS1* gene. (Marshall et al., 2005, 2007b; Astuti et al., 2017).

### Search strategy

The PubMed and Google Scholar databases were searched in January 2022 using the following keyword combination: (“Alström syndrome” OR ALMS) AND (“genotype phenotype” OR “cohort” OR “case report”). The screening of the search results was carried out by BB-M by initially examining the title and abstract to determine whether it corresponded to the subject of the analysis. Some of the articles analysed in this study were not initially found by the database search but were detected by the recommendations and bibliography of other more relevant works. The search was conducted in English only, covering a period between the origin of the databases and the date of the search (January 2022).

### Study selection

All articles selected in a first search were carefully reviewed to meet the criteria for inclusion in the meta-analysis. After a thorough reading of the article, the study was included if the following characteristics were met: the cohort of the article included a patient with a diagnosis of ALMS, and the diagnosis had a genetic and clinical characterisation. Articles whose diagnosis of ALMS was based solely on phenotype were discarded, (Charles et al., 1990; Holder et al., 1995; Russell-Eggitt et al., 1998; Koray et al., 2001; Worthley and Zeitz, 2001; Benso et al., 2002; Satman et al., 2002; Paisey et al., 2004; Hoffman et al., 2005; Hamamy et al., 2006; Koç et al., 2006; Gogi et al., 2007; Silan et al., 2013; Bronson et al., 2015; Boerwinkle et al., 2017; Davies et al., 2018), as well as those that simply presented or reported the patient’s mutations without giving an complete (Lazar et al., 2015) or individualised clinical history (Patel et al., 2006; Marshall et al., 2007b, 2015; Redin et al., 2012; Kilpinen et al., 2017; Gao et al., 2019; Baig et al., 2020). From an initial selection of 108 studies, 31 (Millay et al., 1986; Charles et al., 1990; Holder et al., 1995; Russell-Eggitt et al., 1998; Koray et al., 2001; Worthley and Zeitz, 2001; Benso et al., 2002; Satman et al., 2002; Wu et al., 2003; Iannello et al., 2004; Paisey et al., 2004; Hoffman et al., 2005; Marshall et al., 2005, 2007b, 2015; Koç et al., 2006; Patel et al., 2006; Hamamy et al., 2006; Gogi et al., 2007; Hitz et al., 2008; Catrinoiu et al., 2009; Redin et al., 2012; Silan et al., 2013; Lazar et al., 2015; Bronson et al., 2015; Ahmad et al., 2016; Huang et al., 2016; Lindsey et al., 2017; Boerwinkle et al., 2017; Gao et al., 2019; Baig et al., 2020) were discarded and 76 (Titomanlio et al., 2004; Bond et al., 2005; Minton et al., 2006; Joy et al., 2007; Özgül et al., 2007; Malm et al., 2008; Aldahmesh et al., 2009; Khoo et al., 2009; Liu et al., 2009; Kocova et al., 2011; Wang et al., 2011, 2016a, 2016b, 2021; Aliferis et al., 2012; Jatti et al., 2012; Piñeiro-Gallego et al., 2012; Taokesen et al., 2012; Corbetti et al., 2013; Katagiri et al., 2013; Kuburović et al., 2013; Liang et al., 2013; Mahamid et al., 2013; Taşdemir et al., 2013; Kaya et al., 2014; Bahmad et al., 2014; Louw et al., 2014; Ozantürk et al., 2014; Sanyoura et al., 2014; Casey et al., 2014; Khan et al., 2015; Kim et al., 2015; Long et al., 2015; Sathya Priya et al., 2015; Chakroun et al., 2016; Laxer et al., 2016; Nikopoulos et al., 2016; Xu et al., 2016; Zmyslowska et al., 2016; Chen et al., 2017; Cruz-Aguilar et al., 2017; Das Bhowmik et al., 2017; Hollander et al., 2017; Astuti et al., 2017; Jinda et al., 2017; Bakar et al., 2017; Wicher et al., 2017; Brofferio et al., 2017; Yang et al., 2017; Castro-Sánchez et al., 2017; Dotan et al., 2018; Kilinc et al., 2018; Kvarnung et al., 2018; Maltese et al., 2018; Nasser et al., 2018; Sanchez-Navarro et al., 2018; Tsai et al., 2018; Waldman et al., 2018; Shurygina et al., 2019; Weiss et al., 2019; Gatticchi et al., 2020; Hirano et al., 2020; Hull et al., 2020; Kamal et al., 2020; Lombardo et al., 2020; Mauring et al., 2020; Rethanavelu et al., 2020; Spinelli et al., 2020; Torkamandi et al., 2020; Zhou et al., 2020; Bea-Mascato et al., 2021; Saadah et al., 2021; Srikrupa et al., 2021; Zhang et al., 2021) were selected for information extraction and subsequent analyses.

### Data extraction and curation

For the extraction of information, three main groups were defined: metadata, genetic information, and clinical information.

The metadata refers to the: ID of each patient within the dataset, the ID of the family to which they belong (patients with the same family ID are siblings) and the reference from where the information of that patient has been extracted.

In the case of genetic information, whenever possible, the following should be extracted: the original allele reference, the nomenclature of the mutation for the cDNA and protein sequence, the exon/intron where the mutation is located and the genotype of the patient (homozygote or compound heterozygote). All reported mutations were named following the HGVS guidelines with the transcript NM_015120.4 and validated with the name checker of the mutalyzer software (Lefter et al., 2021). The back translator tool of the mutalyzer software was also used to determine the mutation nomenclature at the cDNA level when only the protein annotation of the mutation was provided in the article. In these cases, only nonsense and stop mutations could be determined, as the algorithm does not work with frameshift mutations.

With regard to the clinical information, sex, age and ethnicity were extracted (whenever possible), the diagnostic criteria named by Marshall et al., (Marshall et al., 2007a) were used to try to homogenise it: vision impairments (history of nystagmus in infancy/childhood, legal blindness, cone and rod dystrophy by ERG); obesity and/or insulin resistance and/or T2DM; history of DCM/CHF; hearing loss; hepatic disfunction; renal failure; pulmonary dysfunction; short stature; males: hypogonadism; females: irregular menses and/or hyperandrogenism; thyroid disorders; mental retarded; abnormal appearance of a finger, Intestinal problems, scoliosis/flat wide feet, epilepsy and alopecia **(Table 1)**. Initially, the full information reported in the article was noted for each individual symptom and the age of onset if this was reported. This was then converted into a binary matrix to simplify and homogenise the subsequent analysis.

**Table 1.** Abbreviations used in the analysis, full name of the phenotypic categories, phenotypes aggregated in each category, prevalence of each phenotypic category in the study cohort and representative HPO terms for each category.

| Abbrev. | Complete name | Phenotype | Prevalence (%) | HPO terms added in each category |
| --- | --- | --- | --- | --- |
| VI | Vision impairments | History of nystagmus in infancy/childhood, legal blindness, cone and rod dystrophy by ERG | 96.54 | HP:0000556 (Retinal dystrophy)\| HP:0000639 (Nystagmus) \|HP:0000618 (Blindness) |
| MT | Metabolic anomalies | Obesity and/or insulin resistance and/or Type 2I diabetes mellitus (T2DM) | 85.88 | HP:0001513 (Obesity)\|HP:0005978 (T2DM) |
| HL | Hearing anomalies | Hearing loss | 62.54 | HP:0000365 (Hearing loss) |
| HRT | Heart anomalies | History of Dilated cardiomyopathy/congestive heart failure (DCM/CHF) | 49.86 | HP:0001644 (DCM)\| HP:0001635 (CHF) |
| LIV | Liver anomalies | Hepatic dysfunction | 34.58 | HP:0001410 (Decreased liver function) |
| REN | Renal anomalies | Renal failure | 29,68 | HP:0000077 (Renal anomaly) |
| MEND | Mental anomalies | Mental disability | 22.48 | HP:0001249 (Intellectual disability) |
| REP | Reproductive system anomalies | Males: hypogonadism; Females: irregular menses and/or hyperandrogenism | 15.56 | HP:0000026 (Male hypogonadism) \|HP:0000858 (Irregular menstruation) |
| PUL | Pulmonary anomalies | Pulmonary dysfunction | 14.99 | HP:0002795 (Impaired pulmonary function) |
| SCO | Spine/feet anomalies | Scoliosis/flat wide feet | 14.12 | HP:0002650 (Scoliosis)\| HP:0001763 (Pes planus) |
| TYD | Thyroid metabolism anomalies | Thyroid disorders | 11.53 | HP:0000820 (Abnormality of the thyroid gland) |
| SHS | Stature anomalies | Short stature | 7.78 | HP:0004322 (Short stature) |
| ALO | Alopecia | Alopecia | 4.61 | HP:0001596 (Alopecia) |
| NER | Nervous system anomalies | Epilepsy | 3.75 | HP:0001250 (Seizure) |
| ABFING | Finger anomalies | Abnormal appearance of a finger | 1.15 | HP:0001155 (Abnormality of the hand) |
| INT | Intestine anomalies | Intestinal problems | 1.15 | HP:0011024 (Abnormality of the gastrointestinal tract) |

### Statistical frequency analysis

To determine the existence of a genotype-phenotype correlation in our patient registry, we selected a sub-cohort of 227 patients who met the following criteria: **I)** complete genetic information (Detection of two mutated alleles with complete notation at the cDNA and protein level), **II)** complete clinical information (Sex, age and presence or absence of the 5 most prevalent clinical manifestations used in these patients: vision impairments, metabolic anomalies, hearing anomalies, heart anomalies and liver anomalies). The heterogeneity of the clinical notation associated with these 5 categories was summarised in a binary matrix (1=presence, 0=absence) which was used for frequency calculations to determine the prevalence of each symptom in the cohort and the different subgroups.

Following the methodology described by Niederlova et al., (Niederlova et al., 2019) a syndromic score between 0 and 1 was calculated. This syndromic score gives an estimate of how many of the 5 most prevalent clinical manifestations described above are present in each patient.

In this study the lack of race was not considered to exclude a patient from being included in the study cohort. Due to the high heterogeneity of this characteristic and the small cohort size, correlations with enough statistical power could not be established.

Patient ages were initially aggregated into 10-year age intervals: 0-9, 10-19, 20-29, 30-39, 40-49, 50-59. After studying the evolution of the syndromic score in the different age intervals, it was found that the number of patients above 39 years was very small. This is consistent with the life expectancy of patients with ALMS, which is usually not more than 50 years. For this reason, a regrouping of the last 3 intervals in patients over 30 years of age was made for the subgroup analysis.

Following the methodology used by Marshall et al., (Marshall et al., 2007b) patients were genotypically grouped according to the different mutational hotspots: exon 8 (E8), exon 10 (E10) and exon 16 (E16). For compound heterozygous patients this aggregation was performed based on the patient’s longest allele. Since ALMS is an autosomal recessive disease, it is expected that a single functional copy of the gene is sufficient to prevent the appearance of the symptoms of the disease (Collin et al., 2002; Hearn, 2018). In this way, it would be expected that a protein truncated in exon 16-20 could have more possibilities of maintaining a certain functionality than if it is truncated in exon 5.

Approximately 90% of the mutations were found in the mutation hotspots (E8, E10, and E16), but to include the remaining % in the study, 3 intervals were defined: longest allele truncated before exon 9 (E8), longest allele truncated between exon 9 and 14 (E10), longest allele truncated after exon 14 (E16).

In cases where only two groups were compared, a Wilcoxon test was used. In trials where comparisons were made between several groups (more than 2), an overall p-value calculation was performed using the non-parametric Kruskal-Wallis test, followed by a comparison of means by groups of two using the Wilcoxon test with correction for FDR, with corrected p-values of less than 0.05 being considered significant.

For the analysis of the prevalence of symptoms in the different genetic groups, contingency tables were created showing the number of positive/negative cases within each patient group. Differences in the prevalence of phenotypes in the different patient groups were assessed using pairwise Fisher’s exact test. Statistical significance of differences between individual groups was determined with the FDR correction for multiple comparisons.

## Results

### Cohort description

A total of 357 patients were collected from a total of 76 scientific publications that passed the initial screening. Only patients with information on sex, age and a complete genetic characterisation (description of two non-functional alleles; **methods**) were used for further analysis. Only mutations in coding regions were considered in the study. This reduced the initial cohort to a total of 227 patients, 128 (56.38%) males and 99 females (43.61%). This study cohort contained a total of 176 variants where 168 were cLOF and 8 missense mutations **(Figure S1)**. Most of them were private mutations, but 5 mutations were shown to have a high frequency: p.(Arg2722Ter) (28 alleles), p.(Gln3495Ter) (22 alleles), p.(Thr3592LysfsTer6) (14 alleles), p.(Glu3773TrpfsTer18) (11 alleles) and p.(Pro3911GlnfsTer16) (10 alleles) **(Figure 1A)**. The number of alleles described for these mutations shows certain inconsistencies with the gnomAD database and in some cases the variant does not even appear in gnomAD database **(Table 2)**.

**Table 2.** Differences in the number of alleles reported in gnomAD and in our database for the most recurrent mutations.

| Pathogenic variant | ALMSDB | gnomAD |
| --- | --- | --- |
| p.(Arg2722Ter)/p.(Arg2720Ter) | 28 | 4 |
| p.(Gln3495Ter)/p.(Gln3493Ter) | 22 | 11 |
| p.(Thr3592LysfsTer6)/p.(Thr3590LysfsTer6) | 14 | 14 |
| p.(Glu3773TrpfsTer18)/p.(Glu3771TrpfsTer18) | 11 | 6 |
| p.(Pro3911GlnfsTer16) | 10 | 0 |

**Figure 1.**
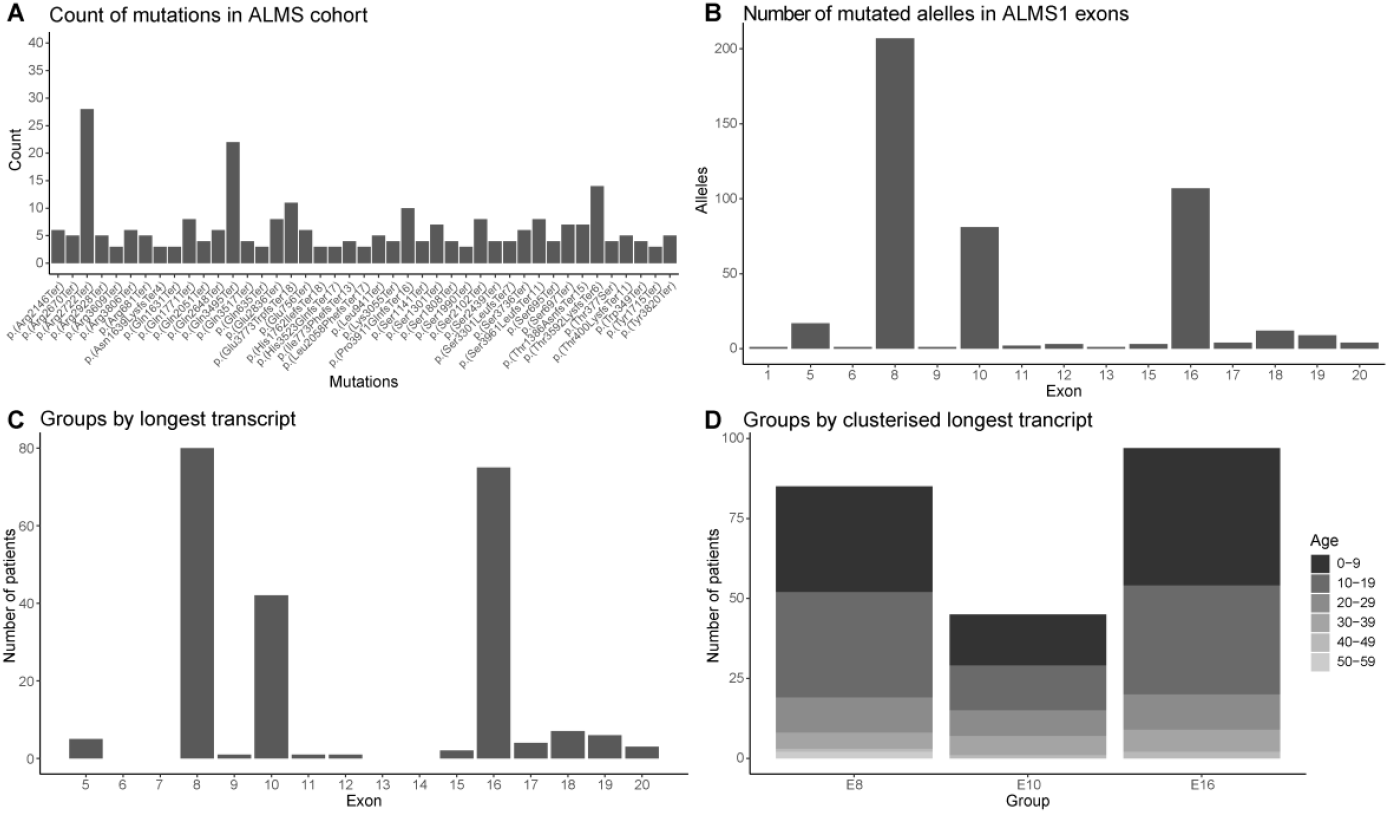
Cohort Description of 227 ALMS patients. **(A)** Counting of the different alleles in the cohort. Alleles with more than 2 copies in the cohort are represented. **(B)** Number of alleles per exon in the study cohort. **(C)** Patients grouped by their copy of the *ALMS1* gene with the later truncation mutation. **(D)** Age composition in the subgroups with the latest truncation site in the *ALMS1* gene in exon 8 (E8), exon 10 (E10) and exon 16 (E16).

Patients were initially grouped into age ranges by decades (from 0 to 59 years) but due to the low number of patients in the age ranges 40-49 and 50-59 years, they were added to the 30-39 years group and reclassified as over 30 years for further analysis.

Out of a total of 454 alleles, 207 (45.59%) were in exon 8, 107 (23.57%) in exon 16 and 81 (17.84%) in exon 10 **(Figure 1B)**. Making these exons the main mutation hotspots in our cohort, consistent with the literature.

Due to the recessive nature of ALMS, we decided to define the characteristic allele of each patient according to the mutation furthest from the origin of the gene **(Figure 1C)**. Many of the patients carrying mutations in exon 8 were compound heterozygotes with mutations in exon 16. After this regrouping we saw that 80 (35.34%) patients had the largest allele truncated in exon 8, 42 (18.50%) in exon 10 and 75 (33.03%) in exon 16.

In accordance with other similar study (Marshall et al., 2007b) we used the mutation hotspots (exons 8, 10 and 16) to subdivide the patients into genetic groups E8, E10 and E16 respectively. Due to the low frequency of mutations in other exons of the *ALMS1* gene we decided to add patients with truncated alleles in adjacent exons to these mutation hotspots. Group E8 includes all patients with the longest truncated allele before exon 9. Group E10 includes patients with the longest truncated allele between exon 9 and exon 13. Finally group E16 included patients with the longest truncated allele from exon 14 to exon 20. As a result, groups E8, E10 and E16 contained 85, 45 and 97 patients respectively **(Figure 1D)**.

### Patients with mutations in exon 10 have a higher syndromic score than the other subgroups

For the strategy of collecting phenotypic manifestations, 16 syndromic groups **(methods)** were initially defined. In the genotype-phenotype correlation analysis, a minimum prevalence threshold of 15% (n=33) was established in the cohort **(Figure S2)**. This reduced the initial phenotypic manifestations to 9 groups: visual impairments (VI, 97.80%), metabolic anomalies (MT, 85.02%), hearing anomalies (HL, 59.91%), heart anomalies (HRT, 49.34%), liver anomalies (LIV, 36.56%), renal anomalies (REN, 29.52%), mental anomalies (MEND, 24.67%), reproductive system anomalies (REP, 17.62%), pulmonary anomalies (RES, 19.38%).

Following the methodology developed by Niederlova et al., (Niederlova et al., 2019), the 5 main disease conditions (VI, MT, HL, HRT and LIV; **methods**) were used to create a discrete syndromic score ranging from 0 to 1 **(Figure 2A)**. The mean of this syndromic score in the total cohort is around 0.7, with the most common values being 0.6 (3 of the 5 symptoms) and 0.8 (4 of the 5 symptoms) **(Figure 2B)**. No significant differences in syndromic score were observed between sexes. However, the syndromic score increased significantly in the different age ranges of the patients (p-value = 3.1 e-10; **Figure 2C-D**). This is consistent with the progressive worsening that these patients undergo throughout their lives (Marshall et al., 2005, 2007a). Finally, we observed how the syndromic score was distributed in the different groups defined according to their largest transcript. Patients whose longest transcript was truncated around exon 10 (E10) had a higher syndromic score compared to the E8 (p-value = 0.017) and E16 (p-value = 0.016) groups **(Figure 2E)**.

**Figure 2.**
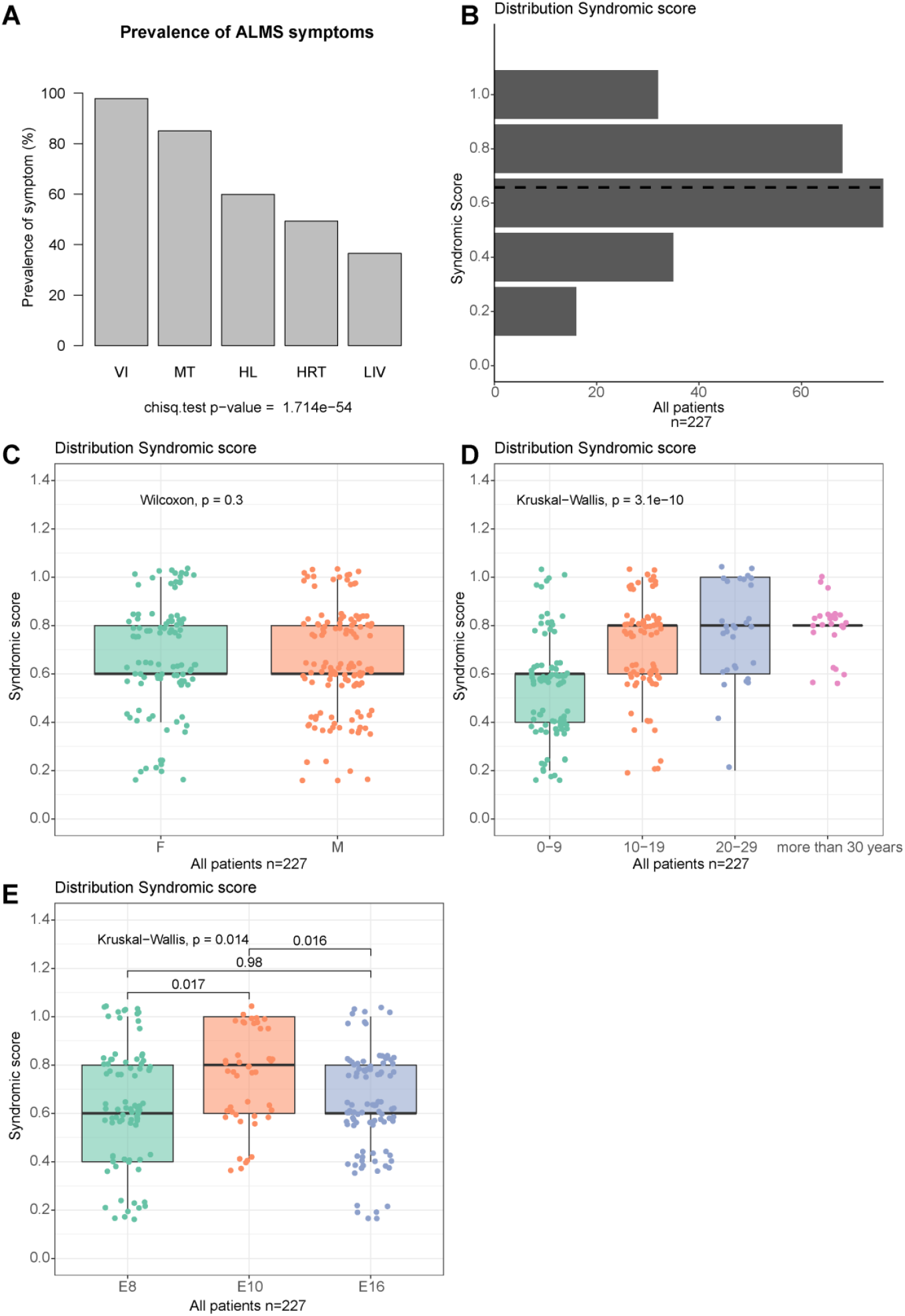
Study of the phenotypic impact on different subgroups of the study cohort. **(A)** Prevalence of the 5 main phenotypic groups in the cohort: visual impairments (VI), metabolic anomalies (MT), hearing anomalies (HL), heart anomalies (HRT) and liver anomalies (LIV). **(B)** Distribution of syndromic scores calculated from the presence or absence of the 5 most relevant phenotypic groups. **(C)** Gender comparison of syndromic scores. **(D)** Comparison of the syndromic scores between age groups. **(E)** Comparison of the syndromic scores between subgroups created from the longest copy of the *ALMS1* gene.

### Patients with truncation variants around exon 10 have a higher prevalence of liver disease

To determine whether the differences between the E8, E10 and E16 groups were due to an unequal composition between the different age ranges in each group, the syndromic score was studied by combining both parameters (age and genetic group) **(Figure 3A)**. In the first decade of life, no significant differences in syndromic score were observed between the groups. Between the ages of 10 to 19 years, a higher syndromic score was observed in the E10 group compared to the E16 group (FDR = 0.061). From 20-29 years of age, a higher syndromic score was observed in the E10 group compared to the E8 and E16 groups (FDR = 0.009, in both cases). Finally, in patients over 30 years of age, no significant differences were observed between the genetic groups.

**Figure 3.**
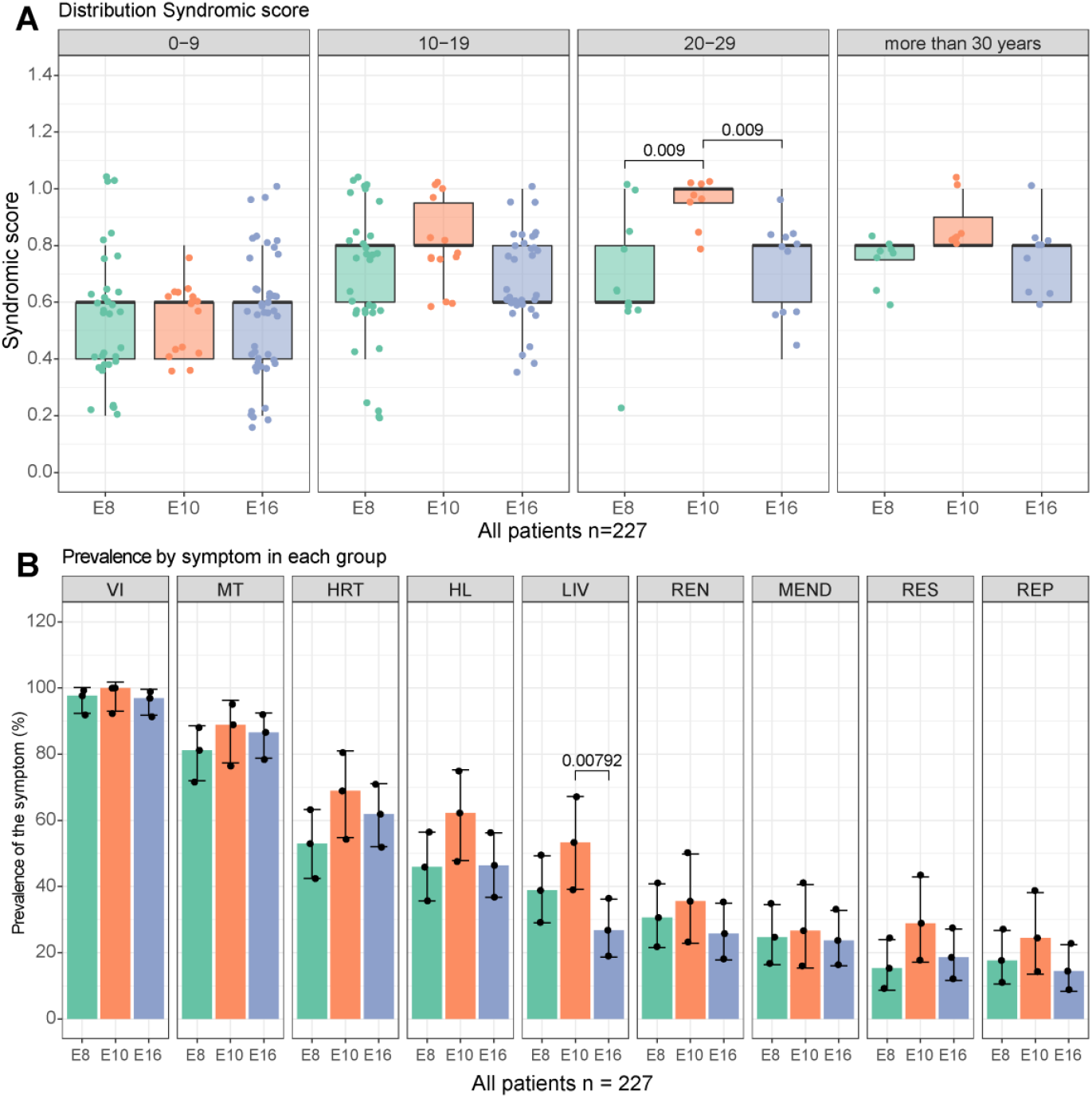
Correlation study between symptoms prevalence and the truncation site of the *ALMS1* gene. **(A)** Evolution of the syndromic score among the subgroups by the longest copy of the *ALMS1* gene in the different age groups. **(B)** Prevalence of the different symptom clusters in the subgroups by the longest copy of the *ALMS1* gene.

After concluding that the differences observed between the groups were not due to an unequal age composition between them, the prevalence of the 9 phenotypic manifestations in each of these groups was studied **(Figure 3B)**. Patients within the E10 group were found to have a higher prevalence of liver disorders compared to patients in the E8 and E16 groups. For the remaining 8 syndromic manifestations no significant differences were found between the genetic groups.

## Discussion

Meta-analyses are a useful tool to address one of the main limitations when attempting to establish a genotype-phenotype correlation in a rare disease, the inability to recruit a large cohort to draw robust and statistically significant conclusions. Although meta-analyses mainly focus on the aggregation of several studies with well-defined and well-studied cohorts, this is often not possible in rare diseases. Alternatively, one can try to aggregate information obtained from case reports in the literature. However, the lack of uniform criteria in how authors describe their patients is one of the main biases of this approach. In this paper we adapt and apply the methodology described by Niederlova et al (Niederlova et al., 2019) from a polygenic disease such as BBS to a monogenic disease such as ALMS.

Despite the fact that ALMS is a monogenic disease (Hearn et al., 2002), to date several studies have already discussed the possible existence of several tissue-specific isoforms for the *ALMS1* gene, which may have different regulatory roles that explain the high symptomatic heterogeneity (Collin et al., 2002, 2012; Hearn et al., 2002; Braune et al., 2017). Although this is an interesting approach, the isolated nature of the data published in the case reports does not allow validation it. In order to validate this hypothesis, the recruitment of a large cohort is necessary. Different cohorts have been described in the literature, the largest being Marshall et al. (Marshall et al., 2007b), followed by Ozantürk et al. (Ozantürk et al., 2014), the NIH clinical centre cohort (Brofferio et al., 2017; Waldman et al., 2018), Chen et al. (Chen et al., 2017) and Rethanavelu et al. (Rethanavelu et al., 2020) with 58, 44, 38, 23 and 21 ALMS patients, respectively. Significant genotype-phenotype correlations were only detected in Marshall et al (Marshall et al., 2007b), highlighting the importance of the sample size used in this type of study.

The causal mutations of ALMS are mainly of the cLOF type (generating a PTC) (Marshall et al., 2015). These types of mutations activate nonsense-mediated decay (NMD), preventing the translation of the sequence from mRNA to protein (Supek et al., 2021). However, certain situations have been described, such as PTC mutations in the last exons of a gene, in which NMD can be prevented or incompletely produced (Lindeboom et al., 2016; Supek et al., 2021). Chen et al., have detected *ALMS1* gene expression in patients carrying two cLOF mutations leading to PTC (Chen et al., 2017). That study has also correlated residual *ALMS1* gene expression with the development of milder phenotypes (Chen et al., 2017). Taking this into account, it could happen that cLOF mutations in the terminal exons of the *ALMS1* gene do not activate NMD, generating a partially functional protein or a non-functional misfolded protein. Under this hypothesis, we grouped the patients in our study according to the longest transcript they had. Three different groups related to the main mutational hotspots of the *ALMS1* gene, exons 8, 10 and 16, were defined. A similar approach has already been used on a cohort of 58 ALMS patients by Marshall et al. (Marshall et al., 2007b). Subsequently, a syndromic score was created by adding the 5 clinical manifestations more prevalent of ALMS. This methodology was adapted from Niederlova et al, (Niederlova et al., 2019).

The results showed that the syndromic score increases with age **(Figure 2D)**, which was consistent with the gradual worsening that these patients suffer throughout their lives (Marshall et al., 2005, 2007a). This helped us to validate the effectiveness of the syndromic score in measuring the severity of the patient symptoms. When comparing the syndromic scores of the different genetic groups by age groups, it was observed that patients with truncation mutations in exon 10 (E10) evolve more unfavourably than the rest **(Figure 3A)**. The fact that these differences are not appreciable after the age of 30 years could be due to a higher mortality in this group in the second and third decades of life. This supposed higher mortality after 30 years could be correlated with the greater prevalence of liver problems that patients with truncation mutations at exon 10 (E10) appear to have **(Figure 3B**). This correlation was not detected in previous genotype-phenotype analyses (Bond et al., 2005; Minton et al., 2006; Patel et al., 2006; Marshall et al., 2007b). Marshall et al.,(Marshall et al., 2007b) described disease-causing variants in exon 16 as leading to urological dysfunction, DCM and diabetes. In our study, we did not look for correlations in urological dysfunction due to the lack of data reported in the literature, leading to a low prevalence of these types of symptoms. In the case of DCM and diabetes the prevalence was the same among the established genetic groups. Marshall et al., (Marshall et al., 2007b) also found a significant association between alterations in exon 8 and absent, mild or delayed kidney disease; findings that were not observed in our analysis.

The explanation for this event could be related to the residual expression of *ALSM1* gene. Mutations in exon 10 could prevent NMD activation and give rise to a misfolded protein that causes greater hepatotoxicity. In the case of mutations in exon 16 or adjacent, perhaps the NMD is not activated either, but the generated protein, despite not being functional, could have a semi-normal conformation that prevents the formation of aggregates. This hypothesis could be easily tested if these protein isoforms can be simulated by homology, but unfortunately the protein structure of the *ALMS1* gene remains unknown and cannot be simulated using the artificial intelligence models such as AlphaFold (van Breugel et al., 2022).

Finally, the *ALMS1* sequence has already described the presence of a lncRNA, *ALMS1-IT1*, with a role in regulating proliferation in various types of cancers and neuroinflammation and a pseudo gene, *ALMS1P1*, whose function is still unknown (Lu et al., 2021; Luan et al., 2021; Mei et al., 2021). Due to the large length of the *ALMS1* gene, the presence of more of these elements would not be uncommon and could explain why the localisation of the cLOF variant can lead to different phenotypes. These events could be of help understanding the regulation of *ALMS1* in the different tissues involved in the disease, and the great heterogeneity observed for the clinical symptoms.

## Conclusion

In conclusion, 5 highly prevalent mutations were detected in our cohort. Not all of these variants appear in public databases. There are no gender differences in the prevalence of the different symptoms of ALMS. Patients’ symptoms worsen with age, and the site of the *ALMS1* gene mutation does not influence the worsening of symptoms in most cases. Only truncation of the ALMS1 protein at exon 10 level correlated with a higher prevalence of liver problems in these patients.

## Data Availability

All data produced are available online at https://github.com/BreisOne/meta-analysis_ALMS and http://genesandfunctions.uvigo.es/alms-db/

http://genesandfunctions.uvigo.es/alms-db/

## Author Contributions

BB-M, DV designed the study. BB-M selected, reviewed, collected and curated data from the scientific articles. BB-M designed and executed the analysis pipeline. BB-M created the publicly accessible online database. BB-M and DV drafted the article. All authors wrote the manuscript and provided approval for publication.

## Informed Consent Statement

Not Apply.

## Data Availability Statement

Data are available via https://github.com/BreisOne/meta-analysis_ALMS or http://genesandfunctions.uvigo.es/alms-db/.

## Code Availability Statement

The code used in this study can be consulted on the GitHub repository https://github.com/BreisOne/meta-analysis_ALMS.

## References

Ahmad, A., D’Souza, B., Yadav, C., Agarwal, A., Kumar, A., Nandini, M., et al. (2016). Metabolic Syndrome in Childhood: Rare Case of Alstrom Syndrome with Blindness. Indian J. Clin. Biochem. 31, 480–482. doi: 10.1007/s12291-015-0543-8.

Aldahmesh, M. A., Abu-Safieh, L., Khan, A. O., Al-Hassnan, Z. N., Shaheen, R., Rajab, M., et al. (2009). Allelic heterogeneity in inbred populations: The Saudi experience with Alström syndrome as an illustrative example. Am. J. Med. Genet. Part A 149, 662– 665. doi: 10.1002/ajmg.a.32753.

Aliferis, K., Hellé, S., Gyapay, G., Duchatelet, S., Stoetzel, C., Mandel, J. L., et al. (2012). Differentiating Alström from Bardet-Biedl syndrome (BBS) using systematic ciliopathy genes sequencing. Ophthalmic Genet. 33, 18–22. doi: 10.3109/13816810.2011.620055.

Astuti, D., Sabir, A., Fulton, P., Zatyka, M., Williams, D., Hardy, C., et al. (2017). Monogenic diabetes syndromes: Locus-specific databases for Alström, Wolfram, and Thiamine-responsive megaloblastic anemia. Hum. Mutat. 38, 764–777. doi: 10.1002/humu.23233.

Bahmad, F., Costa, C. S. A., Teixeira, M. S., de Barros Filho, J., Viana, L. M., and Marshall, J. (2014). Síndrome de Alström familiar: Uma rara causa de perda auditiva progressiva bilateral. Braz. J. Otorhinolaryngol. 80, 99–104. doi: 10.5935/1808-8694.20140023.

Baig, S., Paisey, R., Dawson, C., Barrett, T., Maffei, P., Hodson, J., et al. (2020). Defining renal phenotype in Alström syndrome. Nephrol. Dial. Transplant 35, 994–1001. doi: 10.1093/ndt/gfy293.

Bakar, A. A., Kamal, N. M., Alsaedi, A., Turkistani, R., and Aldosari, D. (2017). Alström syndrome. Med. (United States) 96, 4–7. doi: 10.1097/MD.0000000000006192.

Bea-Mascato, B., Solarat, C., Perea-Romero, I., Jaijo, T., Blanco-Kelly, F., Millán, J. M., et al. (2021). Prevalent alms1 pathogenic variants in spanish alström patients. Genes (Basel). 12, 1–12. doi: 10.3390/genes12020282.

Benso, C., Hadjadj, E., Conrath, J., and Denis, D. (2002). Three new cases of Alström syndrome. Graefe’s Arch. Clin. Exp. Ophthalmol. 240, 622–627. doi: 10.1007/s00417-002-0479-6.

Boerwinkle, C., Marshall, J. D., Bryant, J., Gahl, W. A., Olivier, K. N., and Gunay-Aygun, M. (2017). Respiratory manifestations in 38 patients with Alström syndrome. Pediatr. Pulmonol. 52, 487–493. doi: https://doi.org/10.1002/ppul.23607.

Bond, J., Flintoff, K., Higgins, J., Scott, S., Bennet, C., Parsons, J., et al. (2005). The importance of seeking ALMS1 mutations in infants with dilated cardiomyopathy. J. Med. Genet. 42, 1–3. doi: 10.1136/jmg.2004.026617.

Braune, K., Volkmer, I., and Staege, M. S. (2017). Characterization of Alstrom Syndrome 1 (ALMS1) Transcript Variants in Hodgkin Lymphoma Cells. PLoS One 12, e0170694. Available at: https://doi.org/10.1371/journal.pone.0170694.

Brofferio, A., Sachdev, V., Hannoush, H., Marshall, J. D., Naggert, J. K., Sidenko, S., et al. (2017). Characteristics of cardiomyopathy in Alström syndrome: Prospective singlecenter data on 38 patients. Mol. Genet. Metab. 121, 336–343. doi: 10.1016/J.YMGME.2017.05.017.

Bronson, S. C., Anand Moses, C. R., Periyandavar, I., Dharmarajan, P., Suresh, E., Shanmugam, A., et al. (2015). Diabetes in the young – A case of Alström syndrome with myopathy. J. R. Coll. Physicians Edinb. 45, 33–37. doi: 10.4997/JRCPE.2015.108.

Casey, J., McGettigan, P., Brosnahan, D., Curtis, E., Treacy, E., Ennis, S., et al. (2014). Atypical Alstrom syndrome with novel ALMS1 mutations precluded by current diagnostic criteria. Eur. J. Med. Genet. 57, 55–59. doi: 10.1016/j.ejmg.2014.01.007.

Castro-Sánchez, S., Álvarez-Satta, M., Tohamy, M. A., Beltran, S., Derdak, S., and Valverde, D. (2017). Whole exome sequencing as a diagnostic tool for patients with ciliopathy-like phenotypes. PLoS One 12, 1–16. doi: 10.1371/journal.pone.0183081.

Catrinoiu, D., Mihai, C. M., Tuta, L., Stoicescu, R., and Simpetru, A. (2009). Rare case of Alstrom syndrome with empty sella and interfamilial presence of Bardet-Biedl phenotype. J. Med. Life 2, 98–103.

Chakroun, A., Ben Said, M., Ennouri, A., Achour, I., Mnif, M., Abid, M., et al. (2016). Longterm clinical follow-up and molecular testing for diagnosis of the first Tunisian family with Alström syndrome. Eur. J. Med. Genet. 59, 444–451. doi: 10.1016/J.EJMG.2016.08.004.

Charles, S. J., Moore, A. T., Yates, J. R. W., Green, T., and Clark, P. (1990). Alstrom’s syndrome: Further evidence of autosomal recessive inheritance and endocrinological dysfunction. J. Med. Genet. 27, 590–592. doi: 10.1136/jmg.27.9.590.

Chen, J.-H., Geberhiwot, T., Barrett, T. G., Paisey, R., and Semple, R. K. (2017). Refining genotype-phenotype correlation in Alström syndrome through study of primary human fibroblasts. Mol. Genet. genomic Med. 5, 390–404. doi: 10.1002/mgg3.296.

Collin, G. B., Marshall, J. D., Ikeda, A., So, W. V., Russell-Eggitt, I., Maffei, P., et al. (2002). Mutations in ALMS1 cause obesity, type 2 diabetes and neurosensory degeneration in Alström syndrome. Nat. Genet. 31, 74–8. doi: 10.1038/ng867.

Collin, G. B., Marshall, J. D., King, B. L., Milan, G., Maffei, P., Jagger, D. J., et al. (2012). The Alström syndrome protein, ALMS1, interacts with α-actinin and components of the endosome recycling pathway. PLoS One 7, e37925–e37925. doi: 10.1371/journal.pone.0037925.

Corbetti, F., Razzolini, R., Bettini, V., Marshall, J. D., Naggert, J., Tona, F., et al. (2013). Alström Syndrome: Cardiac Magnetic Resonance findings. Int. J. Cardiol. 167, 1257– 1263. doi: 10.1016/j.ijcard.2012.03.160.

Cruz-Aguilar, M., Galaviz-Hernández, C., Hiebert-Froese, J., Sosa-Macías, M., and Zenteno, J. C. (2017). A nonsense ALMS1 mutation underlies alström syndrome in an extended mennonite kindred settled in north Mexico. Genet. Test. Mol. Biomarkers 21, 397–401. doi: 10.1089/gtmb.2016.0391.

Das Bhowmik, A., Gupta, N., Dalal, A., and Kabra, M. (2017). Whole exome sequencing identifies a homozygous nonsense variation in ALMS1 gene in a patient with syndromic obesity. Obes. Res. Clin. Pract. 11, 241–246. doi: 10.1016/j.orcp.2016.09.004.

Davies, G., Lam, M., Harris, S. E., Trampush, J. W., Luciano, M., Hill, W. D., et al. (2018). Study of 300,486 individuals identifies 148 independent genetic loci influencing general cognitive function. Nat. Commun. 9, 2098. doi: 10.1038/s41467-018-04362-x.

Dotan, G., Khetan, V., Marshall, J. D., Affel, E., Armiger-, D., Naggert, J. K., et al. (2018). Alström syndrome. 38, 440–445. doi: 10.1080/13816810.2016.1257029.Spectraldomain.

Fokkema, I. F. A. C., Taschner, P. E. M., Schaafsma, G. C. P., Celli, J., Laros, J. F. J., and den Dunnen, J. T. (2011). LOVD v.2.0: the next generation in gene variant databases. Hum. Mutat. 32, 557–563. doi: 10.1002/HUMU.21438.

Gao, F. J., Li, J. K., Chen, H., Hu, F. Y., Zhang, S. H., Qi, Y. H., et al. (2019). Genetic and Clinical Findings in a Large Cohort of Chinese Patients with Suspected Retinitis Pigmentosa. Ophthalmology 126, 1549–1556. doi: 10.1016/j.ophtha.2019.04.038.

Gatticchi, L., Miertus, J., Maltese, P. E., Bressan, S., De Antoni, L., Podracká, L., et al. (2020). A very early diagnosis of Alström syndrome by next generation sequencing. BMC Med. Genet. 21, 1–6. doi: 10.1186/s12881-020-01110-1.

Gogi, D., Bond, J., Long, V., Sheridan, E., and Woods, C. G. (2007). Exudative retinopathy in a girl with Alström syndrome due to a novel mutation. Br. J. Ophthalmol. 91, 983– 984. doi: 10.1136/bjo.2005.088781.

Hamamy, H., Barham, M., Alkhawaldeh, A. E. K., Cockburn, D., Snowden, H., and Ajlouni, K. (2006). Alstrom syndrome in four sibs from northern Jordan. Ann. Saudi Med. 26, 480–483. doi: 10.5144/0256-4947.2006.480.

Hearn, T. (2018). ALMS1 and Alström syndrome: a recessive form of metabolic, neurosensory and cardiac deficits. J. Mol. Med. doi: 10.1007/s00109-018-1714-x.

Hearn, T., Renforth, G. L., Spalluto, C., Hanley, N. A., Piper, K., Brickwood, S., et al. (2002). Mutation of ALMS1, a large gene with a tandem repeat encoding 47 amino acids, causes Alström syndrome. Nat. Genet. 31, 79. Available at: https://doi.org/10.1038/ng874.

Hirano, M., Satake, W., Moriyama, N., Saida, K., Okamoto, N., Cha, P. C., et al. (2020). Bardet–Biedl syndrome and related disorders in Japan. J. Hum. Genet. 65, 847–853. doi: 10.1038/s10038-020-0778-y.

Hitz, M. P., Bertram, H., Köditz, H., Görler, H., Happel, C. M., Wessel, A., et al. (2008). Levosimendan for bridging in a pediatric patient with Alström syndrome awaiting heart-lung transplantation. Clin. Res. Cardiol. 97, 846–848. doi: 10.1007/s00392-008-0691-4.

Hoffman, J. D., Jacobson, Z., Young, T. L., Marshall, J. D., and Kaplan, P. (2005). Familial variable expression of dilated cardiomyopathy in Alström syndrome: A report of four sibs. Am. J. Med. Genet. 135 A, 96–98. doi: 10.1002/ajmg.a.30688.

Holder, M., Hecker, W., and Gilli, G. (1995). Impaired glucose tolerance leads to delayed diagnosis of Alström syndrome. Diabetes Care 18, 698–700. doi: 10.2337/diacare.18.5.698.

Hollander, S. A., Alsaleh, N., Ruzhnikov, M., Jensen, K., Rosenthal, D. N., Stevenson, D. A., et al. (2017). Variable clinical course of identical twin neonates with Alström syndrome presenting coincidentally with dilated cardiomyopathy. Am. J. Med. Genet. Part A 173, 1687–1689. doi: 10.1002/ajmg.a.38200.

Huang, L., Xiao, X., Li, S., Jia, X., Wang, P., Sun, W., et al. (2016). Molecular genetics of cone-rod dystrophy in Chinese patients: New data from 61 probands and mutation overview of 163 probands. Exp. Eye Res. 146, 252–258. doi: 10.1016/j.exer.2016.03.015.

Hull, S., Kiray, G., Chiang, J. P. W., and Vincent, A. L. (2020). Molecular and phenotypic investigation of a New Zealand cohort of childhood-onset retinal dystrophy. Am. J. Med. Genet. Part C Semin. Med. Genet. 184, 708–717. doi: 10.1002/ajmg.c.31836.

Iannello, S., Bosco, P., Camuto, M., Cavaleri, A., Milazzo, P., and Belfiore, F. (2004). A mild form of Alstrom disease associated with metabolic syndrome and very high fasting serum afree fatty acids: Two cases diagnosed in adult age. Am. J. Med. Sci. 327, 284–288. doi: 10.1097/00000441-200405000-00031.

Jatti, K., Paisey, R., and More, R. (2012). Coronary artery disease in Alström syndrome. Eur. J. Hum. Genet. 20, 117–118. doi: 10.1038/ejhg.2011.168.

Jinda, W., Taylor, T. D., Suzuki, Y., Thongnoppakhun, W., Limwongse, C., Lertrit, P., et al. (2017). Whole exome sequencing in eight Thai patients with leber congenital amaurosis reveals mutations in the CTNNA1 and CYP4V2 genes. Investig. Ophthalmol. Vis. Sci. 58, 2413–2420. doi: 10.1167/iovs.16-21322.

Joy, T., Cao, H., Black, G., Malik, R., Charlton-Menys, V., Hegele, R. A., et al. (2007). Alstrom syndrome (OMIM 203800): a case report and literature review. Orphanet J. Rare Dis. 2, 49. doi: 10.1186/1750-1172-2-49.

Kamal, N. M., Sahly, A. N., Banaganapalli, B., Rashidi, O. M., Shetty, P. J., Al-Aama, J. Y., et al. (2020). Whole exome sequencing identifies rare biallelic ALMS1 missense and stop gain mutations in familial Alström syndrome patients. Saudi J. Biol. Sci. 27, 271–278. doi: 10.1016/j.sjbs.2019.09.006.

Karczewski, K. J., Francioli, L. C., Tiao, G., Cummings, B. B., Alföldi, J., Wang, Q., et al. (2020). The mutational constraint spectrum quantified from variation in 141,456 humans. Nat. 2020 5817809 581, 434–443. doi: 10.1038/s41586-020-2308-7.

Katagiri, S., Yoshitake, K., Akahori, M., Hayashi, T., Furuno, M., Nishino, J., et al. (2013). Whole-exome sequencing identifies a novel ALMS1 mutation (p.Q2051X) in two Japanese brothers with Alström syndrome. Mol. Vis. 19, 2393–2406. Available at: https://www.ncbi.nlm.nih.gov/pubmed/24319333.

Kaya, A., Orbak, Z., Cąyir, A., Doneray, H., Taşdemir, Ş., Ozanẗurk, A., et al. (2014). Combined occurrence of Alström syndrome and bronchiectasis. Pediatrics 133. doi: 10.1542/peds.2013-0284.

Kerr, T. P., Sewry, C. A., Robb, S. A., and Roberts, R. G. (2001). Long mutant dystrophins and variable phenotypes: evasion of nonsense-mediated decay? Hum. Genet. 109, 402–407. doi: 10.1007/s004390100598.

Khan, A. O., Bifari, I. N., and Bolz, H. J. (2015). Ophthalmic Features of Children Not Yet Diagnosed with Alstrom Syndrome. Ophthalmology 122, 1726–1727.e2. doi: 10.1016/j.ophtha.2015.03.001.

Khoo, E. Y. H., Risley, J., Zaitoun, A. M., El-Sheikh, M., Paisey, R. B., Acheson, A. G., et al. (2009). Alström syndrome and cecal volvulus in 2 siblings. Am. J. Med. Sci. 337, 383– 385. doi: 10.1097/MAJ.0b013e3181926594.

Kilinc, S., Yucel-Yilmaz, D., Ardagil, A., Apaydin, S., Valverde, D., Ozgul, R. K., et al. (2018). Five novel ALMS1 gene mutations in six patients with Alstrom syndrome. J. Pediatr. Endocrinol. Metab. 31, 681–687. doi: 10.1515/jpem-2017-0418.

Kilpinen, H., Goncalves, A., Leha, A., Afzal, V., Alasoo, K., Ashford, S., et al. (2017). Common genetic variation drives molecular heterogeneity in human iPSCs. Nature 546, 370–375. doi: 10.1038/nature22403.

Kim, M. K., Kwak, S. H., Kang, S., Jung, H. S., Cho, Y. M., Kim, S. Y., et al. (2015). Identification of two cases of ciliopathy-associated diabetes and their mutation analysis using whole exome sequencing. Diabetes Metab. J. 39, 439–443. doi: 10.4093/dmj.2015.39.5.439.

Koç, E., Bayrak, G., Suher, M., Ensari, C., Aktas, D., and Ensari, A. (2006). Rare case of Alstrom syndrome without obesity and with short stature, diagnosed in adulthood. Nephrology 11, 81–84. doi: 10.1111/j.1440-1797.2006.00443.x.

Kocova, M., Sukarova-Angelovska, E., Kacarska, R., Maffei, P., Milan, G., and Marshall, J. D. (2011). The unique combination of dermatological and ocular phenotypes in Alström syndrome: severe presentation, early onset and two novel ALMS1 mutations. Br. J. Dermatol. 164, 878–880. doi: 10.1111/j.1365-2133.2010.10157.x.

Koray, F., Dorter, C., Benderli, Y., Satman, I., Yilmaz, T., Dinccag, N., et al. (2001). Alstrom syndrome: a case report. J. Oral Sci. 43, 221–224. doi: 10.2334/josnusd.43.221.

Kuburović, V., Marshall, J. D., Collin, G. B., Nykamp, K., Kuburović, N., Milenković, T., et al. (2013). Differences in the clinical spectrum of two adolescent male patients with Alström syndrome. Clin. Dysmorphol. 22, 7–12. doi: 10.1097/MCD.0b013e32835b9017.

Kvarnung, M., Taylan, F., Nilsson, D., Anderlid, B. M., Malmgren, H., Lagerstedt-Robinson, K., et al. (2018). Genomic screening in rare disorders: New mutations and phenotypes, highlighting ALG14 as a novel cause of severe intellectual disability. Clin. Genet. 94, 528–537. doi: 10.1111/cge.13448.

Landrum, M. J., Lee, J. M., Benson, M., Brown, G. R., Chao, C., Chitipiralla, S., et al. (2018). ClinVar: improving access to variant interpretations and supporting evidence. Nucleic Acids Res. 46, D1062–D1067. doi: 10.1093/NAR/GKX1153.

Laxer, C., Rahman, S. A., Sherif, M., Tahir, S., Cayir, A., Demirbilek, H., et al. (2016). A novel ALMS1 homozygous mutation in two Turkish brothers with Alström syndrome. J. Pediatr. Endocrinol. Metab. 29, 585–589. doi: 10.1515/jpem-2015-0249.

Lazar, C. H., Kimchi, A., Namburi, P., Mutsuddi, M., Zelinger, L., Beryozkin, A., et al. (2015). Nonsyndromic Early-Onset Cone-Rod Dystrophy and Limb-Girdle Muscular Dystrophy in a Consanguineous Israeli Family are Caused by Two Independent yet Linked Mutations in ALMS1 and DYSF. Hum. Mutat. 36, 836–841. doi: 10.1002/humu.22822.

Lefter, M., Vis, J. K., Vermaat, M., den Dunnen, J. T., Taschner, P. E. M., and Laros, J. F. J. (2021). Mutalyzer 2: next generation HGVS nomenclature checker. Bioinformatics 37, 2811–2817. doi: 10.1093/bioinformatics/btab051.

Liang, X., Li, H., Li, H., Xu, F., Dong, F., and Sui, R. (2013). Novel ALMS1 mutations in Chinese patients with alström syndrome. Mol. Vis. 19, 1885–1891.

Lindeboom, R. G. H., Supek, F., and Lehner, B. (2016). The rules and impact of nonsense-mediated mRNA decay in human cancers. Nat. Genet. 2016 4810 48, 1112–1118. doi: 10.1038/ng.3664.

Lindsey, S., Brewer, C., Stakhovskaya, O., Kim, H. J., Zalewski, C., Bryant, J., et al. (2017). Auditory and otologic profile of Alström syndrome: Comprehensive single center data on 38 patients. Am. J. Med. Genet. Part A 173, 2210–2218. doi: 10.1002/ajmg.a.38316.

Liu, L., Dong, B., Chen, X., Li, J., and Li, Y. (2009). Identification of a novel ALMS1 mutation in a Chinese family with Alström syndrome. Eye 23, 1210–1212. doi: 10.1038/eye.2008.235.

Lombardo, B., D’Argenio, V., Monda, E., Vitale, A., Caiazza, M., Sacchetti, L., et al. (2020). Genetic analysis resolves differential diagnosis of a familial syndromic dilated cardiomyopathy: A new case of Alström syndrome. Mol. Genet. Genomic Med. 8, 1–7. doi: 10.1002/mgg3.1260.

Long, P. A., Evans, J. M., Olson, T. M., Gartstein., M. A., Putnam., S., and Kliewer., R. (2015). Exome sequencing establishes diagnosis of Alström syndrome in an infant presenting with non-syndromic dilated cardiomyopathy. Am. J. Med. Genet. A 176, 886–890. doi: 10.1002/ajmg.a.36994.Exome.

Louw, J. J., Corveleyn, A., Jia, Y., Iqbal, S., Boshoff, D., Gewillig, M., et al. (2014). Homozygous loss-of-function mutation in ALMS1 causes the lethal disorder mitogenic cardiomyopathy in two siblings. Eur. J. Med. Genet. 57, 532–535. doi: 10.1016/j.ejmg.2014.06.004.

Lu, P., Zhang, Y., Niu, H., and Wang, Y. (2021). Upregulated Long Non-coding RNA ALMS1-IT1 Promotes Neuroinflammation by Activating NF-κB Signaling in Ischemic Cerebral Injury. Curr. Pharm. Des. 27, 4270–4277. doi: 10.2174/1381612827666210827104316.

Luan, T., Zhang, T. Y., Lv, Z. H., Guan, B. X., Xu, J. Y., Li, J., et al. (2021). The lncRNA ALMS1IT1 may promote malignant progression of lung adenocarcinoma via AVL9-mediated activation of the cyclin-dependent kinase pathway. FEBS Open Bio 11, 1504–1515. doi: 10.1002/2211-5463.13140.

Mahamid, J., Lorber, A., Horovitz, Y., Shalev, S. A., Collin, G. B., Naggert, J. K., et al. (2013). Extreme clinical variability of dilated cardiomyopathy in two siblings with alström syndrome. Pediatr. Cardiol. 34, 455–458. doi: 10.1007/s00246-012-0296-6.

Malm, E., Ponjavic, V., Nishina, P. M., Naggert, J. K., Hinman, E. G., Andréasson, S., et al. (2008). Full-field electroretinography and marked variability in clinical phenotype of Alström syndrome. Arch. Ophthalmol. 126, 51–57. doi: 10.1001/archophthalmol.2007.28.

Maltese, P. E., Iarossi, G., Ziccardi, L., Colombo, L., Buzzonetti, L., Crinò, A., et al. (2018). A Next Generation Sequencing custom gene panel as first line diagnostic tool for atypical cases of syndromic obesity: Application in a case of Alström syndrome. Eur. J. Med. Genet. 61, 79–83. doi: 10.1016/j.ejmg.2017.10.016.

Marshall, J. D., Beck, S., Maffei, P., and Naggert, J. K. (2007a). Alström Syndrome. Eur. J. Hum. Genet. 15, 1193. doi: 10.1038/sj.ejhg.5201933.

Marshall, J. D., Bronson, R. T., Collin, G. B., Nordstrom, A. D., Maffei, P., Paisey, R. B., et al. (2005). New Alström syndrome phenotypes based on the evaluation of 182 cases. Arch. Intern. Med. 165, 675–83. doi: 10.1001/archinte.165.6.675.

Marshall, J. D., Hinman, E. G., Collin, G. B., Beck, S., Cerqueira, R., Maffei, P., et al. (2007b). Spectrum of ALMS1 variants and evaluation of genotype-phenotype correlations in Alström syndrome. Hum. Mutat. 28, 1114–23. doi: 10.1002/humu.20577.

Marshall, J. D., Maffei, P., Collin, G. B., and Naggert, J. K. (2011). Alström syndrome: genetics and clinical overview. Curr. Genomics 12, 225–235. doi: 10.2174/138920211795677912.

Marshall, J. D., Muller, J., Collin, G. B., Milan, G., Kingsmore, S. F., Dinwiddie, D., et al. (2015). Alström Syndrome: Mutation Spectrum of ALMS1. Hum. Mutat. 36, 660– 668. doi: 10.1002/humu.22796.

Mauring, L., Porter, L. F., Pelletier, V., Riehm, A., Leuvrey, A. S., Gouronc, A., et al. (2020). Atypical Retinal Phenotype in a Patient With Alström Syndrome and Biallelic Novel Pathogenic Variants in ALMS1, Including a de novo Variation. Front. Genet. 11. doi: 10.3389/fgene.2020.00938.

Mei, J., Cao, G., He, H., Chang, J., Zhang, B., and Mei, Y. (2021). Effects of long non-coding RNA ALMS1-IT1 on the proliferation and migration of colorectal cancer cells via regulating the expressions of miRNA-889-3p and ATAD2. Cancer Res. Clin. 33, 818– 823. doi: 10.3760/CMA.J.CN115355-20210601-00242.

Millay, R. H., Weleber, R. G., and Heckenlively, J. R. (1986). Ophthalmologic and systemic manifestations of Alström’s disease. Am. J. Ophthalmol. 102, 482–490. doi: 10.1016/0002-9394(86)90078-4.

Minton, J. A. L., Owen, K. R., Ricketts, C. J., Crabtree, N., Shaikh, G., Ehtisham, S., et al. (2006). Syndromic obesity and diabetes: Changes in body composition with age and mutation analysis of ALMS1 in 12 United Kingdom kindreds with Alström syndrome. J. Clin. Endocrinol. Metab. 91, 3110–3116. doi: 10.1210/jc.2005-2633.

Nasser, F., Weisschuh, N., Maffei, P., Milan, G., Heller, C., Zrenner, E., et al. (2018). Ophthalmic features of cone-rod dystrophy caused by pathogenic variants in the ALMS1 gene. Acta Ophthalmol. 96, e445–e454. doi: 10.1111/aos.13612.

Niederlova, V., Modrak, M., Tsyklauri, O., Huranova, M., and Stepanek, O. (2019). Meta-analysis of genotype-phenotype associations in Bardet-Biedl syndrome uncovers differences among causative genes. Hum. Mutat. 40, 2068–2087. doi: 10.1002/HUMU.23862.

Nikopoulos, K., Butt, G. U., Farinelli, P., Mudassar, M., Domènech-Estévez, E., Samara, C., et al. (2016). A large multiexonic genomic deletion within the ALMS1 gene causes Alström syndrome in a consanguineous Pakistani family. Clin. Genet. 89, 510–511. doi: 10.1111/cge.12645.

Ozantürk, A., Marshall, J. D., Collin, G. B., Düzenli, S., Marshall, R. P., Candan, Ş., et al. (2014). The phenotypic and molecular genetic spectrum of Alström syndrome in 44 Turkish kindreds and a literature review of Alström syndrome in Turkey. J. Hum. Genet. 60, 1–9. doi: 10.1038/jhg.2014.85.

Özgül, R. K., Satman, I., Collin, G. B., Hinman, E. G., Marshall, J. D., Kocaman, O., et al. (2007). Molecular analysis and long-term clinical evaluation of three siblings with Alström syndrome. Clin. Genet. 72, 351–356. doi: 10.1111/j.1399-0004.2007.00848.x.

Paisey, R. B., Carey, C. M., Bower, L., Marshall, J., Taylor, P., Maffei, P., et al. (2004). Hypertriglyceridaemia in Alström’s syndrome: Causes and associations in 37 cases. Clin. Endocrinol. (Oxf). 60, 228–231. doi: 10.1111/j.1365-2265.2004.01952.x.

Patel, S., Minton, J. A. L., Weedon, M. N., Frayling, T. M., Ricketts, C., Hitman, G. A., et al. (2006). Common variations in the ALMS1 gene do not contribute to susceptibility to type 2 diabetes in a large white UK population. Diabetologia 49, 1209–1213. doi: 10.1007/s00125-006-0227-2.

Piñeiro-Gallego, T., Cortón, M., Ayuso, C., Baiget, M., and Valverde, D. (2012). Molecular approach in the study of Alström syndrome: analysis of ten Spanish families. Mol. Vis. 18, 1794–1802. Available at: https://www.ncbi.nlm.nih.gov/pubmed/22876109.

Redin, C., Le Gras, S., Mhamdi, O., Geoffroy, V., Stoetzel, C., Vincent, M. C., et al. (2012). Targeted high-throughput sequencing for diagnosis of genetically heterogeneous diseases: Efficient mutation detection in Bardet-Biedl and Alström Syndromes. J. Med. Genet. 49, 502–512. doi: 10.1136/jmedgenet-2012-100875.

Rethanavelu, K., Fung, J. L. F., Chau, J. F. T., Pei, S. L. C., Chung, C. C. Y., Mak, C. C. Y., et al. (2020). Phenotypic and mutational spectrum of 21 Chinese patients with Alström syndrome. Am. J. Med. Genet. Part A 182, 279–288. doi: 10.1002/ajmg.a.61412.

Russell-Eggitt, I. M., Clayton, P. T., Coffey, R., Kriss, A., Taylor, D. S. I., and Taylor, J. F. N. (1998). Alstrom syndrome: Report of 22 cases and literature review. Ophthalmology 105, 1274–1280. doi: 10.1016/S0161-6420(98)97033-6.

Saadah, O. I., Banaganapalli, B., Kamal, N. M., Sahly, A. N., Alsufyani, H. A., Mohammed, A., et al. (2021). Identification of a Rare Exon 19 Skipping Mutation in ALMS1 Gene in Alström Syndrome Patients From Two Unrelated Saudi Families. Front. Pediatr. 9, 1–9. doi: 10.3389/fped.2021.652011.

Sanchez-Navarro, I., R. J. da Silva, L., Blanco-Kelly, F., Zurita, O., Sanchez-Bolivar, N., Villaverde, C., et al. (2018). Combining targeted panel-based resequencing and copy-number variation analysis for the diagnosis of inherited syndromic retinopathies and associated ciliopathies. Sci. Rep. 8, 5285. doi: 10.1038/s41598-018-23520-1.

Sanyoura, M., Woudstra, C., Halaby, G., Baz, P., Senée, V., Guillausseau, P. J., et al. (2014). A novel ALMS1 splice mutation in a non-obese juvenile-onset insulin-dependent syndromic diabetic patient. Eur. J. Hum. Genet. 22, 140–143. doi: 10.1038/ejhg.2013.87.

Sathya Priya, C., Sen, P., Umashankar, V., Gupta, N., Kabra, M., Kumaramanickavel, G., et al. (2015). Mutation spectrum in BBS genes guided by homozygosity mapping in an Indian cohort. Clin. Genet. 87, 161–166. doi: 10.1111/cge.12342.

Satman, I., Yilmaz, M. T., Gürsoy, N., Karşidaǧ, K., Dinççaǧ, N., Ovali, T., et al. (2002). Evaluation of insulin resistant diabetes mellitus in Alström syndrome: A long-term prospective follow-up of three siblings. Diabetes Res. Clin. Pract. 56, 189–196. doi: 10.1016/S0168-8227(02)00004-9.

Sawyer, S. L., Hartley, T., Dyment, D. A., Beaulieu, C. L., Schwartzentruber, J., Smith, A., et al. (2016). Utility of whole-exome sequencing for those near the end of the diagnostic odyssey: time to address gaps in care. Clin. Genet. 89, 275–284. doi: 10.1111/CGE.12654.

Shurygina, M. F., Parker, M. A., Schlechter, C. L., Chen, R., Li, Y., Weleber, R. G., et al. (2019). A case report of two siblings with Alstrom syndrome without hearing loss associated with two new ALMS1 variants. BMC Ophthalmol. 19, 1–9. doi: 10.1186/s12886-019-1259-y.

Silan, F., Gur, S., Kadioglu, L. E., Yalcintepe, S. A., Ukinc, K., Uludag, A., et al. (2013). Characteristic findings of alstrom syndrome with a case report. Open J. Clin. Diagnostics 03, 75–77. doi: 10.4236/ojcd.2013.33014.

Spinelli, V., Girolami, F., Marrone, C., Consigli, V., Iascone, M., Passantino, S., et al. (2020). A rare case of pediatric cardiomyopathy: Alström syndrome identified by gene panel analysis. Clin. Case Reports 8, 3369–3373. doi: 10.1002/ccr3.3327.

Srikrupa, N. N., Sripriya, S., Pavithra, S., Sen, P., Gupta, R., and Mathavan, S. (2021). Whole-exome sequencing identifies two novel ALMS1 mutations in Indian patients with Leber congenital amaurosis. Hum. Genome Var. 8. doi: 10.1038/s41439-021-00143-z.

Supek, F., Lehner, B., and Lindeboom, R. G. H. (2021). To NMD or Not To NMD: Nonsense-Mediated mRNA Decay in Cancer and Other Genetic Diseases. Trends Genet. 37, 657–668. doi: 10.1016/J.TIG.2020.11.002.

Taokesen, M., Collin, G. B., Evsikov, A. V., Güzel, A., Özgül, R. K., Marshall, J. D., et al. (2012). Novel Alu retrotransposon insertion leading to Alström syndrome. Hum. Genet. 131, 407–413. doi: 10.1007/s00439-011-1083-9.

Taşdemir, S., Güzel-Ozantürk, A., Marshall, J., Collin, G., Özgül, R., Narin, N., et al. (2013). Atypical presentation and a novel mutation in ALMS1: implications for clinical and molecular diagnostic strategies for Alström syndrome. Clin. Genet. 83, 96–98. doi: 10.1111/J.1399-0004.2012.01883.X.

Titomanlio, L., De Brasi, D., Buoninconti, A., Sperandeo, M. P., Pepe, A., Andria, G., et al. (2004). Alström syndrome: Intrafamilial phenotypic variability in sibs with a novel nonsense mutation of the ALMS1 gene [3]. Clin. Genet. 65, 156–157. doi: 10.1111/j.0009-9163.2004.00204.x.

Torkamandi, S., Rezaei, S., Mirfakhraei, R., Askari, M., Piltan, S., and Gholami, M. (2020). Whole exome sequencing identified two homozygous ALMS1 mutations in an Iranian family with Alström syndrome. Gene 727, 144228. doi: 10.1016/j.gene.2019.144228.

Tsai, M. C., Yu, H. W., Liu, T., Chou, Y. Y., Chiou, Y. Y., and Chen, P. C. (2018). Rare compound heterozygous frameshift mutations in ALMS1 gene identified through exome sequencing in a Taiwanese patient with Alström syndrome. Front. Genet. 9, 1–5. doi: 10.3389/fgene.2018.00110.

van Breugel, M., Rosa e Silva, I., and Andreeva, A. (2022). Structural validation and assessment of AlphaFold2 predictions for centrosomal and centriolar proteins and their complexes. Commun. Biol. 5, 1–10. doi: 10.1038/s42003-022-03269-0.

Waldman, M., Han, J. C., Reyes-Capo, D. P., Bryant, J., Carson, K. A., Turkbey, B., et al. (2018). Alström syndrome: Renal findings in correlation with obesity, insulin resistance, dyslipidemia and cardiomyopathy in 38 patients prospectively evaluated at the NIH clinical center. Mol. Genet. Metab. 125, 181–191. doi: 10.1016/j.ymgme.2018.07.010.

Wang, C., Luo, X., Wang, Y., Liu, Z., Wu, S., Wang, S., et al. (2021). Novel mutations of the ALMS1 gene in patients with alström syndrome. Intern. Med. 60, 3721–3728. doi: 10.2169/internalmedicine.6467-20.

Wang, S., Zhang, Q., Zhang, X., Wang, Z., and Zhao, P. (2016a). Clinical and genetic characteristics of Leber congenital amaurosis with novel mutations in known genes based on a Chinese eastern coast Han population. Graefe’s Arch. Clin. Exp. Ophthalmol. 254, 2227–2238. doi: 10.1007/s00417-016-3428-5.

Wang, X., Feng, Y., Li, J., Zhang, W., Wang, J., Lewis, R. A., et al. (2016b). Retinal diseases caused by mutations in genes not specifically associated with the clinical diagnosis. PLoS One 11, 1–12. doi: 10.1371/journal.pone.0165405.

Wang, X., Wang, H., Cao, M., Li, Z., Chen, X., Patenia, C., et al. (2011). Whole-exome sequencing identifies ALMS1, IQCB1, CNGA3, and MYO7A mutations in patients with Leber congenital amaurosis. Hum. Mutat. 32, 1450–1459. doi: 10.1002/humu.21587.

Weiss, S., Cohen, L., Ben-Yosef, T., Ehrenberg, M., and Goldenberg-Cohen, N. (2019). Late diagnosis of Alstrom syndrome in a Yemenite-Jewish child. Ophthalmic Genet. 40, 7–11. doi: 10.1080/13816810.2018.1561900.

Wicher, K., Bajon, T., Wawrocka, A., Skorczyk-Werner, A., Niedziela, M., and Krawczynski, M. R. (2017). Alström syndrome: A case report of the Polish family and a brief review of the differential diagnosis. Pediatr. Pol. 92, 781–785. doi: 10.1016/j.pepo.2017.07.003.

Worthley, M. I., and Zeitz, C. J. (2001). Case of Alstrmsyndrome with late presentation dilated cardiomyopathy. Intern. Med. J. 31, 569–570. doi: 10.1046/j.1445-5994.2001.00140.x.

Wu, W. C., Chen, S. C., Dia, C. Y., Yu, M. L., Hsieh, M. Y., Lin, Z. Y., et al. (2003). Alström syndrome with acute pancreatitis: A case report. Kaohsiung J. Med. Sci. 19, 358– 360. doi: 10.1016/s1607-551x(09)70438-3.

Xu, Y., Guan, L., Xiao, X., Zhang, J., Li, S., Jiang, H., et al. (2016). ALMS1 null mutations: A common cause of Leber congenital amaurosis and early-onset severe cone-rod dystrophy. Clin. Genet. 89, 442–447. doi: 10.1111/cge.12617.

Yang, L., Li, Z., Mei, M., Fan, X., Zhan, G., Wang, H., et al. (2017). Whole genome sequencing identifies a novel ALMS1 gene mutation in two Chinese siblings with Alström syndrome. BMC Med. Genet. 18, 1–6. doi: 10.1186/s12881-017-0418-3.

Zhang, J. J., Wang, J. Q., Sun, M. Q., Xu, D., Xiao, Y., Lu, W. L., et al. (2021). Alström syndrome with a novel mutation of ALMS1 and Graves’ hyperthyroidism: A case report and review of the literature. World J. Clin. Cases 9, 3200–3211. doi: 10.12998/wjcc.v9.i13.3200.

Zhou, C., Xiao, Y., Xie, H., Liu, S., and Wang, J. (2020). A novel variant in ALMS1 in a patient with Alström syndrome and prenatal diagnosis for the fetus in the family: A case report and literature review. Mol. Med. Rep. 22, 3271–3276. doi: 10.3892/mmr.2020.11398.

Zmyslowska, A., Borowiec, M., Antosik, K., Ploski, R., Ciechanowska, M., Iwaniszewska, B., et al. (2016). Genetic evaluation of patients with Alström syndrome in the Polish population. Clin. Genet. 89, 448–453. doi: 10.1111/cge.12656.

