## Supplementary material for "Meta-analysis of genotype-phenotype associations in Alström syndrome"

*Correspondence: Diana Valverde

### Supplementary Material:

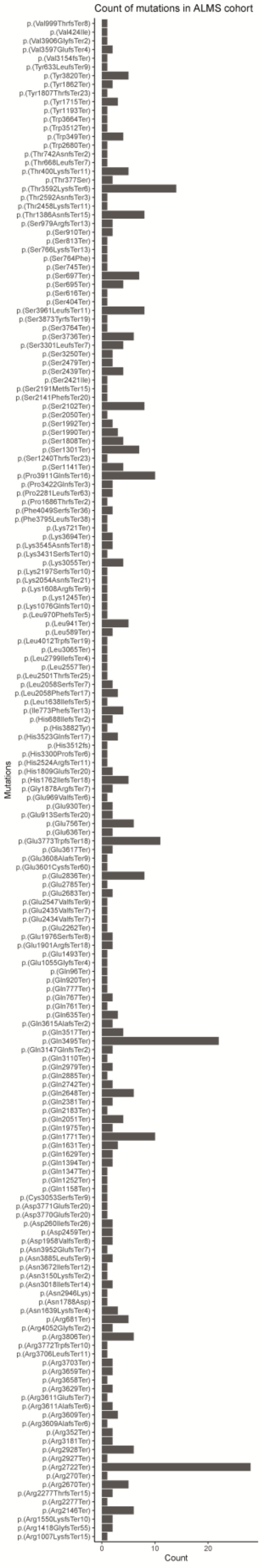

**Supplementary Figure S1.** Graphical representation of the 176 variants and the number of alleles of each variant in the cohort.

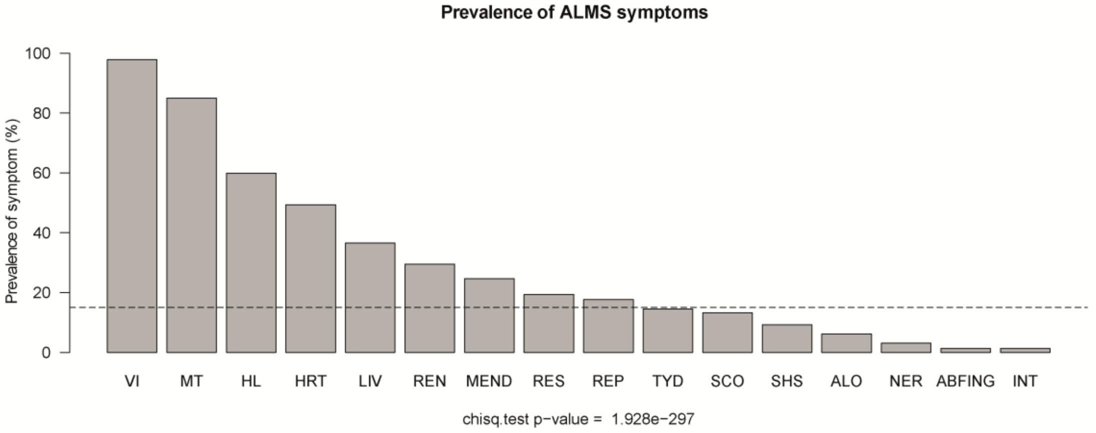


**Supplementary Figure S2.** Graphical representation of the prevalence in the 16 syndromic groups initially collected from the literature and the minimum prevalence limit established (15%; n=33) to be able to be introduced in the study.
